# Uptake of COVID-19 vaccines among pregnant women: a systematic review and meta-analysis

**DOI:** 10.1101/2022.04.01.22273296

**Authors:** Petros Galanis, Irene Vraka, Olga Siskou, Olympia Konstantakopoulou, Aglaia Katsiroumpa, Daphne Kaitelidou

## Abstract

**Background:** Mass vaccination against the COVID-19 is essential to control the pandemic. COVID-19 vaccines are recommended now during pregnancy to prevent adverse outcomes.

**Objective:** To evaluate the evidence from the literature regarding the uptake of COVID-19 vaccination among pregnant women.

**Methods:** We conducted a systematic review following the Preferred Reporting Items for Systematic Reviews and Meta-Analysis guidelines. We searched PubMed, Medline, Scopus, ProQuest, Web of Science, CINAHL, and a pre-print service (medRxiv) from inception to March 23, 2022. We included quantitative studies reporting COVID-19 vaccination uptake among pregnant women, studies that examine predictors of COVID-19 vaccination uptake and studies that examine reasons for decline of vaccination. We performed meta-analysis to estimate the overall proportion of vaccinated pregnant women against the COVID-19.

**Results:** We found 11 studies including 703,004 pregnant women. The overall proportion of vaccinated pregnant women against the COVID-19 was 27.5% (95% CI: 18.8-37.0%). The pooled proportion for studies that were conducted in Israel was higher than the proportion for studies that were conducted in USA and other countries. Predictors of COVID-19 vaccination uptake were older age, ethnicity, race, trust in COVID-19 vaccines, and fear of COVID-19 during pregnancy. On the other hand, mistrust in the government, diagnosis with COVID-19 during pregnancy, and worry about the safety and the side effects of the COVID-19 vaccines were reasons for decline of vaccination.

**Conclusions:** The global COVID-19 vaccination prevalence in pregnant women is low. There is a large gap in the literature on the factors influencing the decision of pregnant women to be vaccinated against the COVID-19. Targeted information campaigns are essential to improve trust and build vaccine literacy among pregnant women. Given the ongoing high case rates and the known increased risks of COVID-19 in pregnant women, our findings could help policy makers to improve the acceptance rate of COVID-19 vaccines in pregnant women especially in vulnerable subgroups.

## Introduction

Pregnant women with COVID-19 are at increased risk for severe illness, adverse birth outcomes and mortality. In particular, hospitalized pregnant women with symptomatic COVID-19 were more likely to have iatrogenic preterm births, to be admitted to intensive care and to need invasive ventilation than pregnant women without COVID-19 (Khalil et al., 2020; Lokken et al., 2021; Vousden et al., 2021; Woodworth et al., 2020; Zambrano et al., 2020). For instance, in United Kingdom, between February and September 2021, 98% of the 1714 pregnant women admitted to hospital with symptomatic COVID-19 were unvaccinated (Iacobucci, 2021), while no fully vaccinated pregnant women were admitted to intensive care with COVID-19 (UK Health Security Agency, 2021).

Pregnant women were not included in the initial randomized controlled trials testing COVID-19 vaccines, leading to the lack of data on vaccination safety and pregnancy outcomes compared to the general population (Pogue et al., 2020; Polack et al., 2020). However, two systematic reviews found that reactogenicity is similar in pregnant women and the general population, abortion rate is similar in vaccinated and non-vaccinated pregnant women studied before the COVID-19 pandemic, and anti-SARS-CoV-2 immunoglobulins are transferred through the placenta and the breast milk to the newborns, providing protective immunity (Falsaperla et al., 2021; Fu et al., 2022). Moreover, according to a systematic review with studies in the USA, pregnant women have the same risk of adverse pregnancy or neonatal outcomes with unvaccinated pregnant women (Rawal et al., 2022). In general, COVID-19 vaccination produces immune responses during pregnancy and does not cause major negative outcomes. Thus, several organizations, such as the Center for Disease Control, the Society for Maternal-Fetal Medicine and the American College of Obstetricians and Gynecologists recommend now that pregnant women should receive COVID-19 vaccines to prevent severe maternal morbidity and adverse birth outcomes (American College of Obstetricians and Gynecologist (ACOG), 2021; Centers for Disease Control and Prevention, 2021; Rasmussen & Jamieson, 2021).

To the best of our knowledge, none of the systematic reviews provided evidence about the uptake of COVID-19 vaccines among pregnant women. Therefore, the aim of this systematic review was to identify what is known about the uptake of COVID-19 vaccines among pregnant women. Also, we investigated predictors of COVID-19 vaccination uptake among pregnant women and reasons for decline of vaccination.

## Methods

### Data sources and strategy

We conducted a systematic review following the Preferred Reporting Items for Systematic Reviews and Meta-Analysis (PRISMA) guidelines (Moher et al., 2009). We searched PubMed, Medline, Scopus, ProQuest, Web of Science, CINAHL, and a pre-print service (medRxiv) from inception to March 23, 2022. We used the following strategy in all fields: ((pregnan*) AND (vaccin*)) AND (covid-19).

### Selection and eligibility criteria

Three independent authors applied a three-step procedure for studies selection: removal of duplicates, screening of title and abstract, and reading of full-text articles. In particular, two independent authors performed study selection and a third, senior author resolved the differences. Moreover, we examined reference lists of all relevant articles. The population of interest was pregnant women and the outcome was the COVID-19 vaccination uptake. Thus, we included quantitative studies reporting COVID-19 vaccination uptake among pregnant women, studies that examine predictors of COVID-19 vaccination uptake and studies that examine reasons for decline of vaccination. We included any paper with information about COVID-vaccination uptake in pregnant women independently the semester of pregnancy. Studies published in English were eligible to be included. We excluded reviews, protocols, posters, case reports, statements, letters to the Editor, expert opinions, and editorials.

### Data extraction and risk of bias assessment

Three reviewers independently extracted the following data from the studies: authors, country, data collection time, sample size, age of pregnant women, study design, sampling method, response rate, percentage of COVID-19 vaccination uptake among pregnant women, predictors of COVID-19 vaccination uptake, reasons for decline of COVID-19 vaccination, and type of publication (journal or pre-print service).

We used the Joanna Briggs Institute critical appraisal tool to assess risk of bias of studies (Santos et al., 2018). The response options are the following: Yes when the criteria are clearly identifiable through the article; No when the criteria are not identifiable; Unclear when the criteria are not clearly identified in the article; and Not applicable when the criteria do not apply to the study. The risk of bias is ranked as “low”, “moderate”, and “high” according to the percentage of “Yes” responses.

### Statistical analysis

The outcome variable was the COVID-19 vaccination uptake among pregnant women. We divided the number of vaccinated pregnant women by the total number of pregnant women to calculate the proportion of pregnant women that took a COVID-19 vaccine. Then, we transformed this proportion with the Freeman-Tukey Double Arcsine method and we calculated the respective 95% confidence intervals (CI) for the proportions (Barendregt et al., 2013). We used I^2^ and the Hedges Q statistics to assess heterogeneity between studies. I^2^ value higher than 75% indicates high heterogeneity, and a p-value<0.1 for the Hedges Q statistic indicates statistically significant heterogeneity (Higgins, 2003). Heterogeneity between results was very high; thus we applied a random effect model to estimate pooled proportion of COVID-19 vaccinated pregnant women (Higgins, 2003). We considered country, data collection time, sample size, age of pregnant women, study design, sampling method, response rate, risk of bias, and publication type (journal or pre-print service) as pre-specified sources of heterogeneity. Due to the scarce data and the high heterogeneity in the results in some variables (e.g. age of pregnant women), we decided to perform subgroup analysis for risk of bias, study design, and the country that studies were conducted. Also, we performed meta-regression analysis using sample size and data collection time as the independent variables. We treated data collection time as a continuous variable giving the number 1 for studies that were conducted in December 2020, the number 2 for studies that were conducted in January 2020, etc. We performed a leave-one-out sensitivity analysis to estimate the influence of each study on the overall proportion of COVID-19 vaccinated pregnant women. We used the funnel plot and the Egger’s test to assess the publication bias. Regarding the Egger’s test, a P-value<0.05 indicating publication bias (Egger et al., 1997). We did not perform meta-analysis for the factors that affect pregnant women’ decision to be vaccinated against the COVID-19 since the data was very scarce. We used OpenMeta[Analyst] for the meta-analysis (Wallace et al., 2009).

## Results

### Identification and selection of studies

Flowchart of our systematic review is shown in Figure 1. Our initial search yielded 6,932 records after duplicates removal. Applying the inclusion and exclusion criteria, we identified 11 articles.

**Figure 1.**
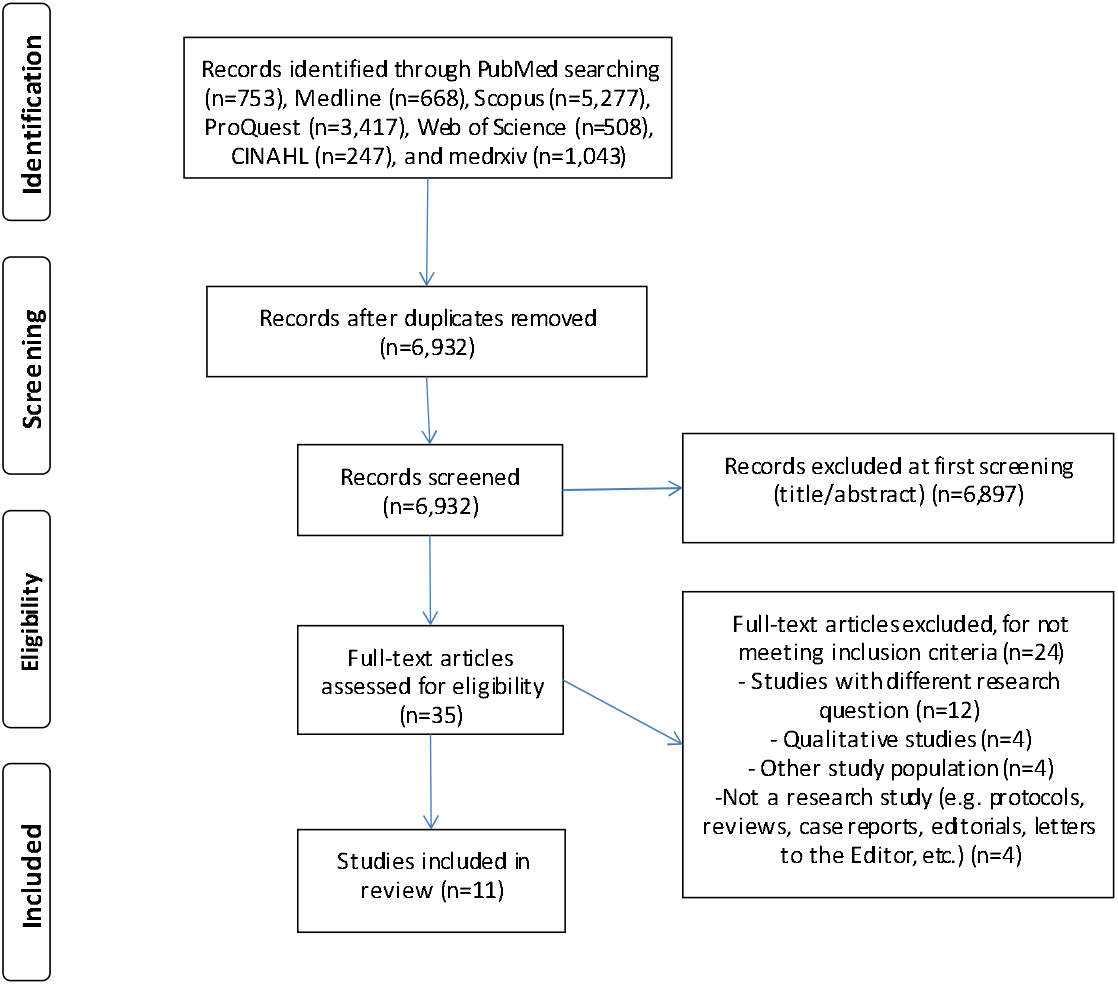
Flowchart of the literature search according to the Preferred Reporting Items for Systematic Reviews and Meta-Analysis.

### Characteristics of the studies

We found 11 studies including 703,004 pregnant women. Main characteristics of the studies included in this review are presented in Table 1. Four studies were conducted in Israel (Goldshtein et al., 2022; Rottenstreich et al., 2022; Taubman – Ben=Ari et al., 2022; Wainstock et al., 2021), three studies in the USA (Lipkind et al., 2022; Razzaghi et al., 2021; Siegel et al., 2021), two studies in United Kingdom (Blakeway et al., 2022; UK Health Security Agency, 2021), one study in Japan (Hosokawa et al., 2022), and one study in Scotland (Stock et al., 2022). Data collection time among studies ranged from December 2020 (Lipkind et al., 2022; Razzaghi et al., 2021) to October 2021 (Stock et al., 2022). Sample size ranged from 473 (Siegel et al., 2021) to 355,299 pregnant women (UK Health Security Agency, 2021). Eight studies were cohort studies (Blakeway et al., 2022; Goldshtein et al., 2022; Lipkind et al., 2022; Razzaghi et al., 2021; Rottenstreich et al., 2022; Stock et al., 2022; UK Health Security Agency, 2021; Wainstock et al., 2021) and three studies were cross-sectional (Hosokawa et al., 2022; Siegel et al., 2021; Taubman – Ben=Ari et al., 2022). Two studies used national data (Stock et al., 2022; UK Health Security Agency, 2021), three studies used a convenience sample (Hosokawa et al., 2022; Siegel et al., 2021; Taubman – Ben=Ari et al., 2022), and six studies did not report the sampling method (Blakeway et al., 2022; Goldshtein et al., 2022; Lipkind et al., 2022; Razzaghi et al., 2021; Rottenstreich et al., 2022; Wainstock et al., 2021). Ten studies were published in peer-reviewed journals (Blakeway et al., 2022; Goldshtein et al., 2022; Hosokawa et al., 2022; Lipkind et al., 2022; Razzaghi et al., 2021; Rottenstreich et al., 2022; Stock et al., 2022; Taubman – Ben=Ari et al., 2022; UK Health Security Agency, 2021; Wainstock et al., 2021) and one study in a pre-print service (Siegel et al., 2021).

**Table 1.** Overview of the studies included in this systematic review.

| Reference | Country | Data collection time | Sample size (N) | Age, mean (standard deviation) | Study design | Sampling method | Response rate (%) | COVID-19 vaccination uptake, % (n/N) | Publication in |
| --- | --- | --- | --- | --- | --- | --- | --- | --- | --- |
| (Hosokawa et al., 2022) Hosokawa | Japan | July 28 to August 30, 2021 | 1,621 | <29 years, 35.6%; ≥29 years, 64.4% | Cross-sectional | Convenience sampling | 73.9 | 13.4 (217/1,621) | Journal |
| (Rottenstreich et al., 2022) Rottenstreich | Israel | January 19 to April 27, 2021 | 1,775 | 30.6 (5.8) for vaccinated and 29.5 (6) for unvaccinated | Cohort | NR | NR | 40.2 (712/1,775) | Journal |
| (Taubman – Ben-Ari et al., 2022) Taubman | Israel | March to April, 2021 | 860 | 28.3 (4.4) | Cross-sectional | Convenience sampling | 65 | 45.2 (389/860) | Journal |
| (Blakeway et al., 2022) Blakeway | United Kingdom | March to July, 2021 | 491 | 35 (NR) for vaccinated and 33 (NR) for unvaccinated | Cohort | NR | NR | 28.5 (140/491) | Journal |
| (Wainstock et al., 2021) Wainstock | Israel | January to June, 2021 | 4,399 | 30.6 (5.3) for vaccinated and 28.2 (5.7) for unvaccinated | Cohort | NR | NR | 20.8 (913/4,399) | Journal |
| (Razzaghi et al., 2021) Razzaghi | USA | December 14, 2020 to May 8, 2021 | 135,968 | 18-24 years, 13.9%; 25-34 years, 61.3%; 35-49 years, 24.8% | Cohort | NR | NR | 16.3 (22,163/135,968) | Journal |
| (Lipkind et al., 2022) Lipkind | USA | December 15, 2020 to July 22, 2021 | 46,079 | 32.3 (4.5) for vaccinated and 29.8 (5.3) for unvaccinated | Cohort | NR | NR | 21.8 (10,064/46,079) | Journal |
| (Stock et al., 2022) Stock | Scotland | December 1, 2020 to October 31, 2021 | 131,751 | NR | Cohort | National data | NA | 9.5 (12,518/131,751) | Journal |
| (UK Health Security | United Kingdom | January to August | 355,299 | NR | Cohort | National | NA | 7 | Journal |
| Agency, 2021) UK Health Security Agency |  | 2021 |  |  |  | data |  | (24,759/355,299) |  |
| (Goldshtein et al., 2022) Goldshtein | Israel | March to September 2021 | 24,288 | 31.6 (5.2) for vaccinated and 30.5 (5.7) for unvaccinated | Cohort | NR | NR | 68.7 (16,697/24,288) | Journal |
| (Siegel et al., 2021) Siegel | USA | June to August 2021 | 473 | 33 (4.5) for vaccinated and 31.4 (5.6) for unvaccinated | Cross-sectional | Convenience sampling | 69.7 | 49.3 (233/473) | Pre-print service |

NA: not applicable; NR: not reported

### Risk of bias assessment

Risk of bias assessment of studies included in this review is shown in Supplementary Table 1. Risk of bias was moderate in five cohort studies (Goldshtein et al., 2022; Lipkind et al., 2022; Rottenstreich et al., 2022; Stock et al., 2022; Wainstock et al., 2021) and low in three cohort studies (Blakeway et al., 2022; Razzaghi et al., 2021; UK Health Security Agency, 2021). The most common bias in cohort studies was the absence of strategies to address incomplete follow up. Also, only one cohort study (Blakeway et al., 2022) used multivariate analysis to eliminate confounding. Regarding cross-sectional studies, risk of bias was low in two studies (Hosokawa et al., 2022; Siegel et al., 2021) and moderate in one study (Taubman – Ben=Ari et al., 2022).

### COVID-19 vaccination uptake

The overall proportion of vaccinated pregnant women against the COVID-19 was 27.5% (95% CI: 18.8-37.0%) (Figure 2). COVID-19 vaccination uptake among pregnant women ranged from 7.0% (95% CI: 6.9-7.1%) (UK Health Security Agency, 2021) to 68.7% (95% CI: 68.2-69.3%) (Goldshtein et al., 2022). The heterogeneity between results was very high (I^2^=99.98%, p-value for the Hedges Q statistic<0.001). A leave-one-out sensitivity analysis showed that no single study had a disproportional effect on the overall proportion, which varied between 23.5% (95% CI: 18.5-28.8%), with Goldshtein et al. (2022) excluded, and 29.7% (95% CI: 18.9-41.9%), with UK Health Security Agency (2021) excluded. Publication bias was probable according to Egger’s test (<0.05) and funnel plot (Supplementary Figure 1).

**Figure 2.**
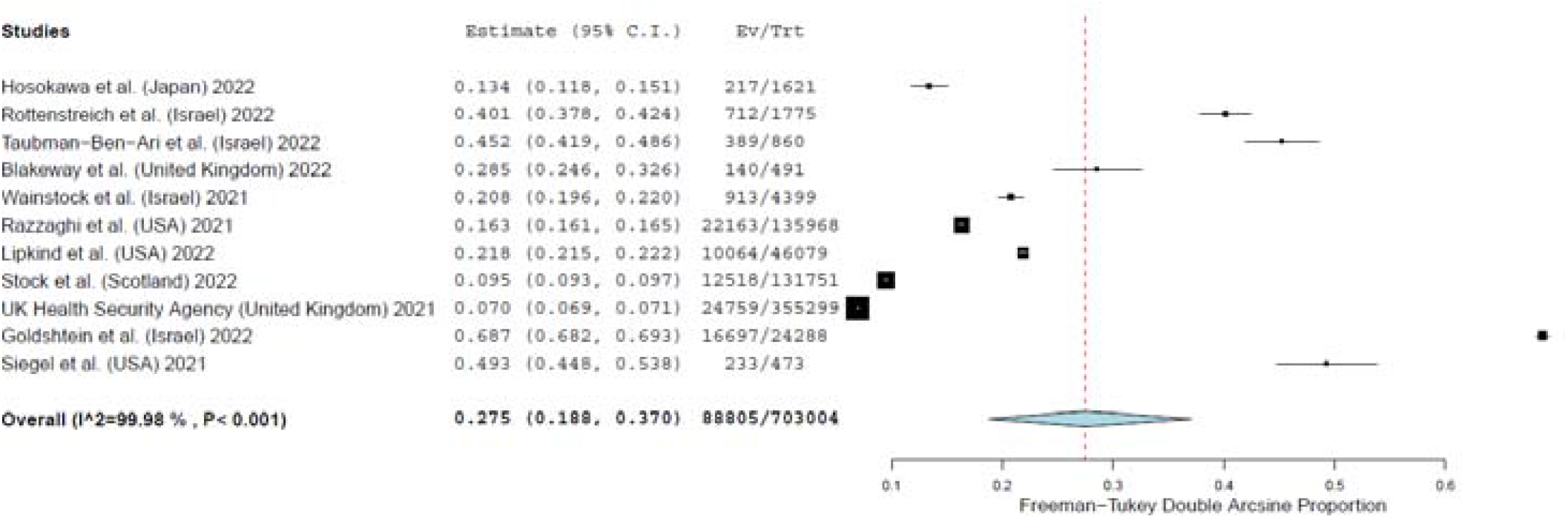
Forest plot of vaccinated pregnant women against the COVID-19.

According to subgroup analysis, the pooled proportion for the studies with moderate risk of bias (33.0% [95% CI: 13.8-55.8%], I^2^=99.99) was higher than the proportion for the studies with low risk of bias (21.0% [95% CI: 13.8-29.3%], I^2^=99.96). Type of study was another source of heterogeneity, since the pooled proportion for the cross-sectional studies (34.7% [95% CI: 11.9-62.1%], I^2^=99.52) was higher than the proportion for the cohort studies (24.9% [95% CI: 15.3-35.9%], I^2^=99.99). Moreover, the pooled proportion for studies that were conducted in Israel (43.3% [95% CI: 17.1-71.8%], I^2^=99.93) was higher than the proportion for studies that were conducted in USA (27.3% [95% CI: 21.6-33.3%], I^2^=99.78) and other countries (12.8% [95% CI: 10.4-15.4%], I^2^=99.71). According to meta-regression analysis, COVID-19 vaccination uptake among pregnant women was independent sample size (p=0.07) and data collection time (p=0.34).

### Factors related with COVID-19 vaccination uptake

Predictors of COVID-19 vaccination uptake among pregnant women and reasons for decline of vaccination are shown in Table 2. Five studies investigated factors that affect pregnant women’ decision to vaccinate against the COVID-19 (Blakeway et al., 2022; Hosokawa et al., 2022; Razzaghi et al., 2021; Siegel et al., 2021; UK Health Security Agency, 2021). Three studies (Blakeway et al., 2022; Hosokawa et al., 2022; Siegel et al., 2021) used multivariate analysis to eliminate confounding, and two studies (Razzaghi et al., 2021; UK Health Security Agency, 2021) used descriptive statistics to present relationships between factors and COVID-19 vaccination uptake among pregnant women.

**Table 2.** Predictors of COVID-19 vaccination uptake among pregnant women and reasons for decline of vaccination.

| Reference | Predictors of COVID-19 vaccination uptake | Reasons for decline of COVID-19 vaccination |
| --- | --- | --- |
| (Blakeway et al., 2022) Blakeway | - Pregestational diabetes mellitus (OR=10.5; 95% CI=1.74 to 83.2; p-value=0.014) |  |
| (Hosokawa et al., 2022) Hosokawa |  | - Mistrust in the government (OR=1.26; 95% CI=1.03 to 1.54; p-value=0.001) |
| (Razzaghi et al., 2021) Razzaghi | - Increased age (35-49 years, 22.7%; 25-34 years, 71.8%; 18-24 years, 5.5%)<br>- Vaccination rate was highest among Asian women (24.7%) and White women (19.7%) and lowest among Black women (6%) and Hispanic women (11.9%) |  |
| (UK Health Security Agency, 2021) UK Health Security | - Increased age ($\leq 24$ years, 7.5% of women were vaccinated; 25-34 years, 27%; 35-44 years, 44.7%; $\geq 45$ years, 22.1%)<br>- Vaccination rate was highest among White women (17.5%) and Asian women (13.5%) and lowest among Black women (5.5%)<br>- Vaccination rate was highest among women lived in least deprived areas (26.5%) and lowest among women lived in most deprived areas (7.8%) | |
| (Siegel et al., 2021) Siegel | - Trust in COVID-19 vaccines (OR=6.5; 95% CI=4.3 to 9.9; p-value<0.05)<br>- Trust in COVID-19 vaccines effectiveness for women (OR=10.8; 95% CI=6.7 to 17.2; p-value<0.05)<br>- Trust in COVID-19 vaccines effectiveness for newborns (OR=6.4; 95% CI=4.2 to 9.7; p-value<0.05)<br>- Fear of COVID-19 during pregnancy (OR=2.5; 95% CI=1.7 to 3.6; p-value<0.05) | - Worry about the safety of the COVID-19 vaccines (OR=0.16; 95% CI=0.10 to 0.27; p-value<0.05)<br>- Worry about the side effects of the COVID-19 vaccines for women (OR=0.18; 95% CI=0.12 to 0.27; p-value<0.05)<br>- Worry about the side effects of the COVID-19 vaccines for newborns (OR=0.17; 95% CI=0.11 to 0.25; p-value<0.05)<br>- Diagnosis with COVID-19 during pregnancy (OR=0.27; 95% CI=0.11 to 0.69; p-value<0.05) |

Two studies (Razzaghi et al., 2021; UK Health Security Agency, 2021) found that increased age was related with increased probability of COVID-19 vaccination uptake. Also, two studies (Razzaghi et al., 2021; UK Health Security Agency, 2021) found that White women and Asian women were vaccinated for COVID-19 more often than Black women and Hispanic women, while one study (UK Health Security Agency, 2021) found that vaccination was highest among women lived in least deprived areas and lowest among women lived in most deprived areas. Trust in COVID-19 vaccines, fear of COVID-19 during pregnancy and pregestational diabetes mellitus were predictors of COVID-19 vaccination uptake among pregnant women (Blakeway et al., 2022; Siegel et al., 2021), while mistrust in the government, diagnosis with COVID-19 during pregnancy, and worry about the safety and the side effects of the COVID-19 vaccines were reasons for decline of vaccination (Hosokawa et al., 2022; Siegel et al., 2021).

## Discussion

To the best of our knowledge, this is the first systematic review and meta-analysis that estimates the COVID-19 vaccination uptake among pregnant women and examines predictors of uptake and reasons for decline of vaccination. Eleven studies met our inclusion and exclusion criteria and we found that worldwide the uptake prevalence of the vaccination against the COVID-19 is 27.5% in pregnant women. This prevalence is considerably lower than the prevalence of pregnant women who expressed the intention to be vaccinated against the COVID-19. In particular, two meta-analyses (Carbone et al., 2022; Nikpour et al., 2022) found that the global prevalence of pregnant women accepting the COVID-19 is about 49-54%. Moreover, in a survey with 5,282 pregnant women from 16 countries, 52% of them indicated an intention to receive a COVID-19 vaccine (Skjefte et al., 2021).

Our review provides evidence of low levels of vaccine uptake in pregnant women. The situation is getting worse since the proportion of pregnant women that actually take a COVID-19 vaccine is even lower in studies with low risk of bias and in the cohort studies. Thus, our estimation is probably an overestimation of the true global prevalence of pregnant women accepting the COVID-19 since the quality and the type of study seems to have a significant impact on the results of the studies. Moreover, we found that the vaccination rate is much higher in Israel than in other countries. This great difference may be due to the fact that Israel was one of the first countries that launched a national vaccination project encouraging all pregnant women to receive a COVID-19 vaccine (The Israeli Health Ministry, 2021). It is notable that four about of the 11 studies included in this review were conducted in Israel. This fact further demonstrates the urgency in Israel to inoculate the entire adult population, including pregnant women, as quickly as possible.

Interestingly, the vaccine uptake rate did not improve even when data from studies have begun to demonstrate the safety and efficacy of COVID-19 vaccines in pregnant women (Shimabukuro et al., 2021; Trostle et al., 2021). However, the number of studies carried out since the publication of this information is very small and not sufficient to draw firm conclusions.

Five out of the 11 studies in this review examined factors that are associated with COVID-19 vaccine uptake in pregnant women. Older age of pregnant women is associated with vaccine uptake. This finding is confirmed by studies that investigated the intention of pregnant women to accept a COVID-19 vaccine. Several studies found that older age is related with higher acceptance of COVID-19 vaccines (Levy et al., 2021; Skjefte et al., 2021; Stuckelberger et al., 2021). This finding is plausible since it is well known that pregnancy at advanced maternal age is a risk factor for adverse outcomes, such as higher rate of neonatal intensive care unit admission, preterm deliveries, spontaneous miscarriage, pre-eclampsia, low birthweight babies, preterm labor, worse Apgar scores, and cesarean deliveries (Glick et al., 2021; Pinheiro et al., 2019). Moreover, older age is associated with higher COVID-19 mortality (Mehraeen et al., 2020; Sepandi et al., 2020; Yanez et al., 2020). It is probable that older pregnant women confront COVID-19 with more fear resulting on a higher COVID-19 vaccination uptake (Skjefte et al., 2021; Tao et al., 2021).

According to our review, COVID-19 vaccination rate was highest among White and Asian pregnant women, and lowest among Black and Hispanic pregnant women. Hispanic ethnicity and Black or African American race is related with refusal of COVID-19 vaccination in pregnancy (Battarbee et al., 2022; Huddleston et al., 2022; Levy et al., 2021; Razzaghi et al., 2021; Townsel et al., 2021). A systematic review found that white individuals have a higher rate of COVID-19 vaccine uptake than black individuals (Galanis et al., 2021). Also, similar racial and ethnic disparities have been reported for the acceptance of other recommended vaccinates in pregnancy, such as tetanus, influenza and acellular pertussis, with Black and Hispanic women have the lowest vaccination coverage (Razzaghi et al., 2020).

Trust in COVID-19 vaccines and fewer worries about the safety and the side effects of the COVID-19 vaccines are predictors of COVID-19 vaccination uptake. Similar factors such as trust in the safety and efficacy of the COVID-19 vaccines, confidence in received information on COVID-19 vaccination, no fear of the COVID-19 vaccines side effects, trust in childhood vaccines, and influenza vaccination within the previous year are associated with a higher intention rate of pregnant women to take a COVID-19 vaccine (Ceulemans et al., 2021; Gencer et al., 2021; Geoghegan et al., 2021; Mappa et al., 2021; Skjefte et al., 2021). In general, high levels of information and knowledge about COVID-19 vaccines decrease fear and have significant influence on the pregnant women decision to undergo COVID-19 vaccination.

### Limitations

Our review and meta-analysis is subject to some limitations. Data taken from databases may not provide the most up-to-date evidence regarding COVID-19 vaccination uptake among pregnant women due to publication process. This limitation is of particular importance in the present review, as the data on vaccination of pregnant women are constantly increasing. Moreover, data collection time among studies ranged from December 2020 to October 2021, while evidence regarding safety and efficacy of COVID-19 vaccines in pregnant women is increasing significantly on an ongoing basis. Thus, we should interpret the results of this review with care since they may not directly predict future behavior of pregnant women. Additionally, we could not generalize our results since the number of relevant studies included in this review is low and these studies were conducted only in five countries.

Only five studies examined the factors that affect pregnant women decision to take a COVID-19 vaccine. Moreover, these studies investigated mainly demographic factors, e.g. age, ethnicity, race, etc. Τhere is a large gap in the literature on the factors influencing the decision of pregnant women to be vaccinated against the COVID-19. For instance, psychological factors and social media variables that could affect women’ attitudes towards COVID-19 vaccination uptake are not investigated so far.

Regarding meta-analysis, we applied a random effects model and we performed subgroup and meta-regression analysis to overcome the high level of the statistical heterogeneity. However limited number of studies, high heterogeneity in the results in some variables, and scarce data forced us to perform subgroup and meta-regression analysis for a few variables. At least, the leave-one-out sensitivity analysis confirmed the robustness of our results.

## Conclusions

We found that the global COVID-19 vaccination prevalence in pregnant women is low. Given the ongoing high case rates and the known increased risks of COVID-19 in pregnant women, high vaccination rate in this vulnerable population is paramount to reduce adverse outcomes, morbidity and mortality. An understanding of the factors related with increased COVID-19 vaccine uptake in pregnant women is essential to improve trust and build vaccine literacy. Moreover, there is a need for different public health messages and targeted information campaigns to improve COVID-19 vaccination acceptance especially in minority groups. Policy makers and healthcare professionals should reduce the fear and anxiety of pregnant women regarding the safety and efficacy of the COVID-19 vaccines. Education about the COVID-19 vaccines with strong and more informative messages is important to increase the acceptance of a COVID-19 vaccine in pregnant women.

## Data Availability

All data produced in the present study are available upon reasonable request to the authors

---

**Supplement Table 1a.**
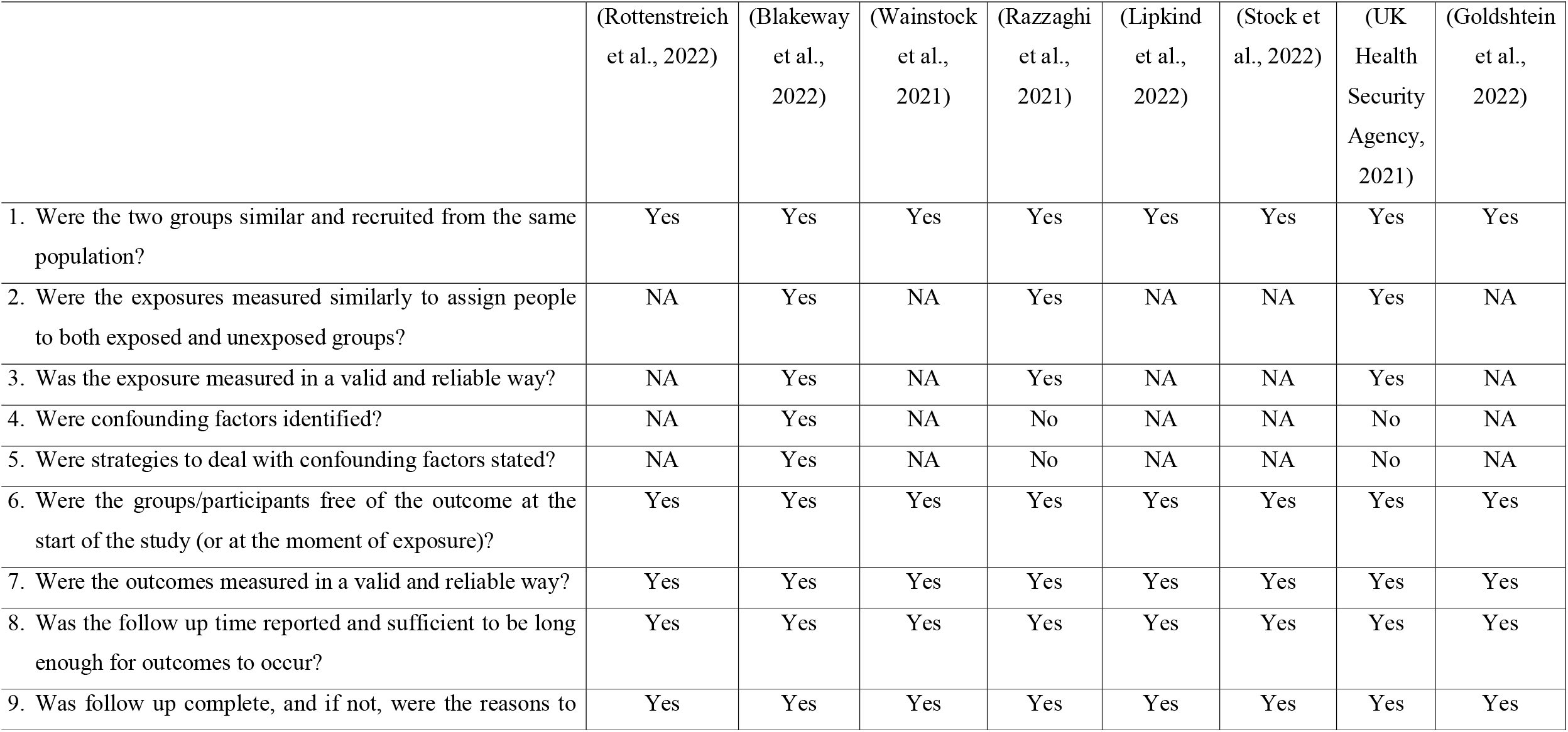
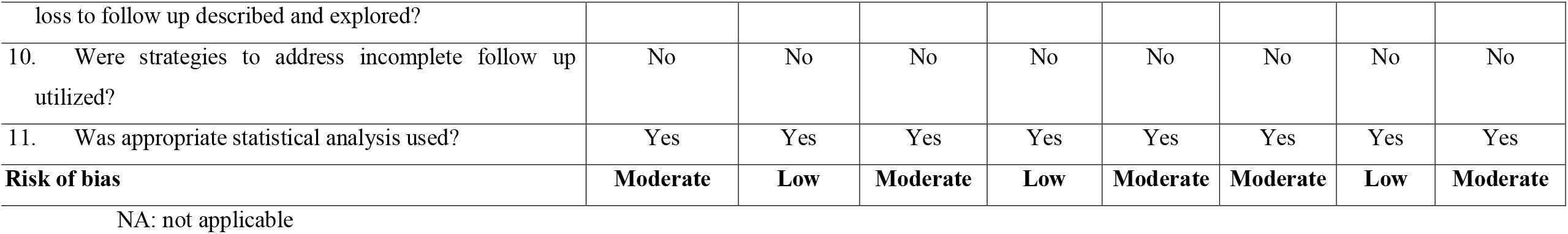
Quality of cohort studies included in this systematic review.

**Supplement Table 1b.**
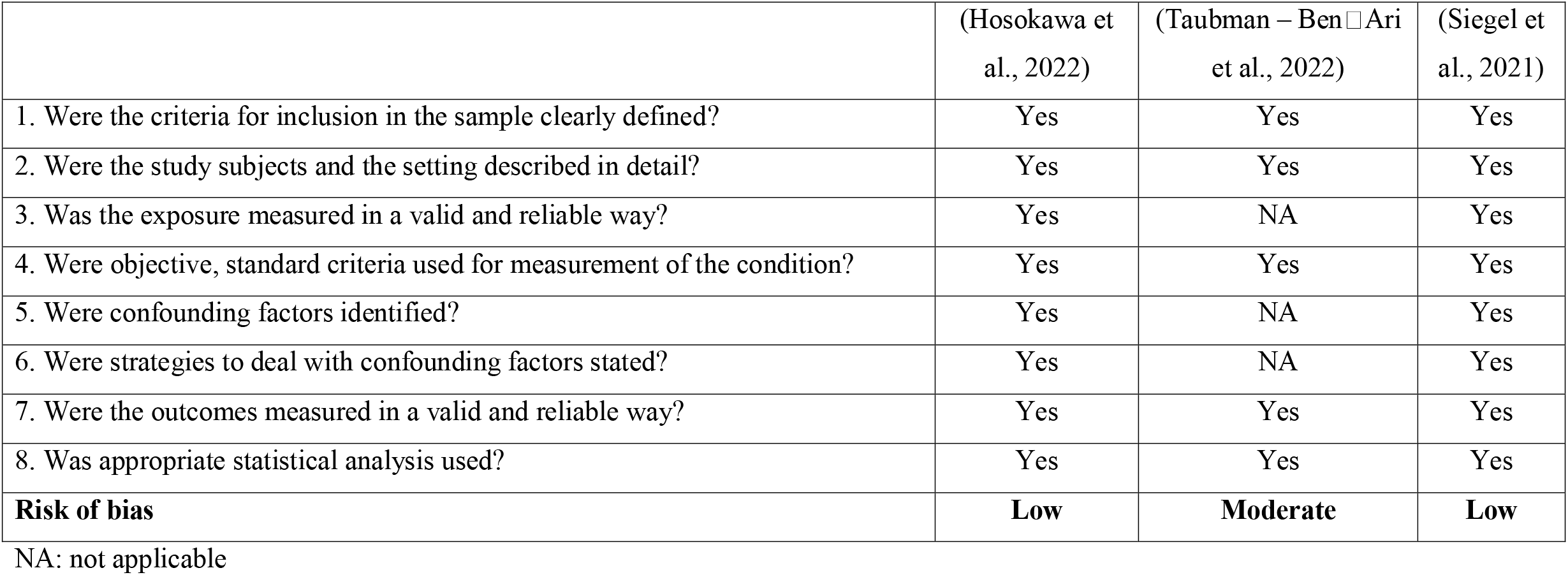
Quality of cross-sectional studies included in this systematic review.

|  | (Hosokawa et al., 2022) | (Taubman – Ben Ari et al., 2022) | (Siegel et al., 2021) |
| --- | --- | --- | --- |
| 1. Were the criteria for inclusion in the sample clearly defined? | Yes | Yes | Yes |
| 2. Were the study subjects and the setting described in detail? | Yes | Yes | Yes |
| 3. Was the exposure measured in a valid and reliable way? | Yes | NA | Yes |
| 4. Were objective, standard criteria used for measurement of the condition? | Yes | Yes | Yes |
| 5. Were confounding factors identified? | Yes | NA | Yes |
| 6. Were strategies to deal with confounding factors stated? | Yes | NA | Yes |
| 7. Were the outcomes measured in a valid and reliable way? | Yes | Yes | Yes |
| 8. Was appropriate statistical analysis used? | Yes | Yes | Yes |
| **Risk of bias** | **Low** | **Moderate** | **Low** |

NA: not applicable

**Supplementary Figure 1.**
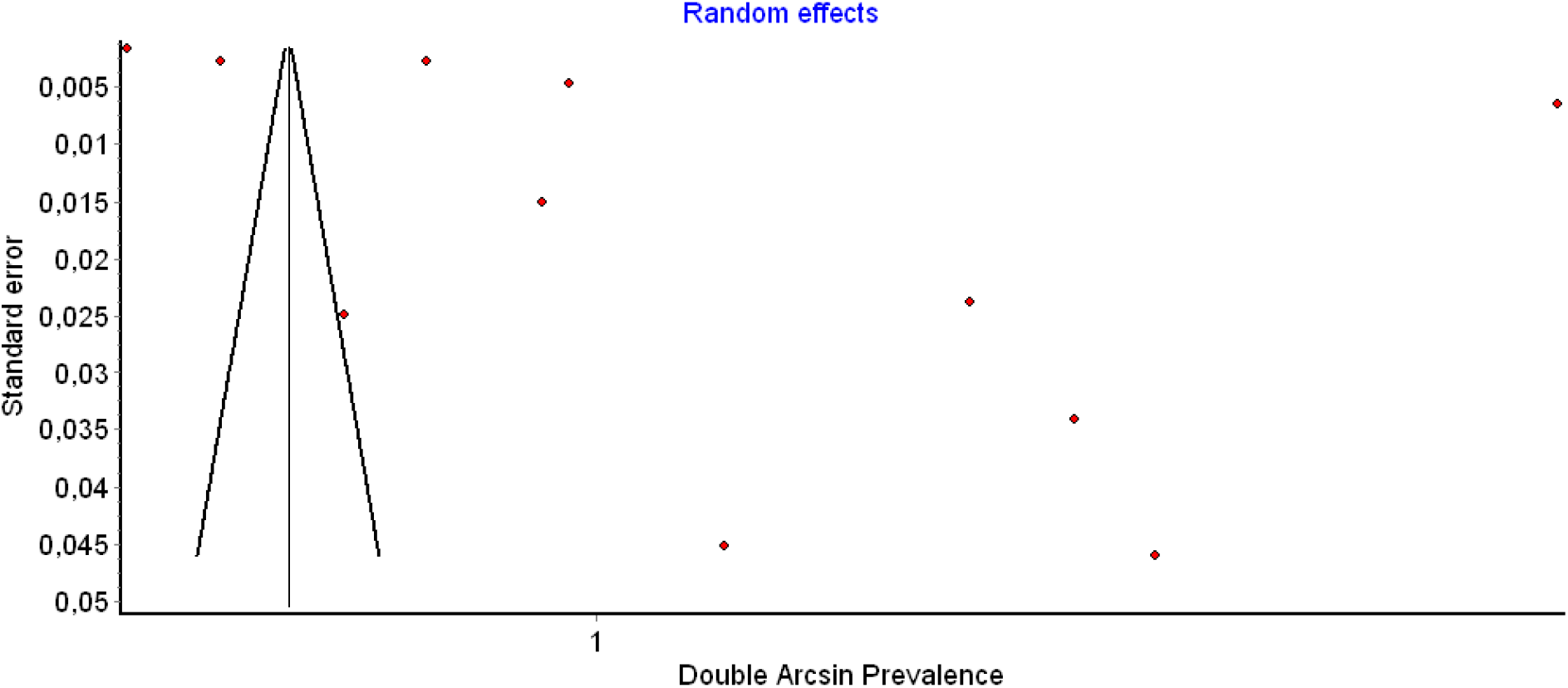
Funnel plot of vaccinated pregnant women against the COVID-19.

## References

American College of Obstetricians and Gynecologist (ACOG). (2021). ACOG and SMFM Recommend COVID-19 Vaccination for Pregnant Individuals. https://www.acog.org/news/newsreleases/2021/07/acog-smfm-recommend-covid-19-vaccination-for-pregnant-individuals

Barendregt, J. J., Doi, S. A., Lee, Y. Y., Norman, R. E., & Vos, T. (2013). Meta-analysis of prevalence. Journal of Epidemiology and Community Health, 67(11), 974–978. https://doi.org/10.1136/jech-2013-203104

Battarbee, A. N., Stockwell, M. S., Varner, M., Newes-Adeyi, G., Daugherty, M., Gyamfi-Bannerman, C., Tita, A. T., Vorwaller, K., Vargas, C., Subramaniam, A., Reichle, L., Galang, R. R., Powers, E., Lucca-Susana, M., Parks, M., Chen, T. J., Razzaghi, H., & Dawood, F. S. (2022). Attitudes Toward COVID-19 Illness and COVID-19 Vaccination among Pregnant Women: A Cross-Sectional Multicenter Study during August–December 2020. American Journal of Perinatology, 39(01), 075–083. https://doi.org/10.1055/s-0041-1735878

Blakeway, H., Prasad, S., Kalafat, E., Heath, P. T., Ladhani, S. N., Le Doare, K., Magee, L. A., O’Brien, P., Rezvani, A., von Dadelszen, P., & Khalil, A. (2022). COVID-19 vaccination during pregnancy: Coverage and safety. American Journal of Obstetrics and Gynecology, 226(2), 236.e1-236.e14. https://doi.org/10.1016/j.ajog.2021.08.007

Carbone, L., Di Girolamo, R., Mappa, I., Saccone, G., Raffone, A., Di Mascio, D., De Vivo, V., D’Antonio, F., Guida, M., Rizzo, G., & Maria Maruotti, G. (2022). Worldwide beliefs among pregnant women on SARS-CoV-2 vaccine: A systematic review. European Journal of Obstetrics & Gynecology and Reproductive Biology, 268, 144–164. https://doi.org/10.1016/j.ejogrb.2021.12.003

Centers for Disease Control and Prevention. (2021). COVID-19 vaccines while pregnant or breastfeeding. https://www.cdc.gov/coronavirus/2019-ncov/vaccines/recommendations/pregnancy.html

Ceulemans, M., Foulon, V., Panchaud, A., Winterfeld, U., Pomar, L., Lambelet, V., Cleary, B., O’Shaughnessy, F., Passier, A., Richardson, J. L., Allegaert, K., & Nordeng, H. (2021). Vaccine Willingness and Impact of the COVID-19 Pandemic on Women’s Perinatal Experiences and Practices—A Multinational, Cross-Sectional Study Covering the First Wave of the Pandemic. International Journal of Environmental Research and Public Health, 18(7), 3367. https://doi.org/10.3390/ijerph18073367

Egger, M., Smith, G. D., Schneider, M., & Minder, C. (1997). Bias in meta-analysis detected by a simple, graphical test. BMJ, 315(7109), 629–634. https://doi.org/10.1136/bmj.315.7109.629

Falsaperla, R., Leone, G., Familiari, M., & Ruggieri, M. (2021). COVID-19 vaccination in pregnant and lactating women: A systematic review. Expert Review of Vaccines, 20(12), 1619–1628. https://doi.org/10.1080/14760584.2021.1986390

Fu, W., Sivajohan, B., McClymont, E., Albert, A., Elwood, C., Ogilvie, G., & Money, D. (2022). Systematic review of the safety, immunogenicity, and effectiveness of COVID-19 vaccines in pregnant and lactating individuals and their infants. International Journal of Gynaecology and Obstetrics: The Official Organ of the International Federation of Gynaecology and Obstetrics, 156(3), 406–417. https://doi.org/10.1002/ijgo.14008

Galanis, P., Vraka, I., Siskou, O., Konstantakopoulou, O., Katsiroumpa, A., & Kaitelidou, D. (2021). Predictors of COVID-19 vaccination uptake and reasons for decline of vaccination: A systematic review [Preprint]. Public and Global Health. https://doi.org/10.1101/2021.07.28.21261261

Gencer, H., Özkan, S., Vardar, O., & Serçekuş, P. (2021). The effects of the COVID 19 pandemic on vaccine decisions in pregnant women. Women and Birth, S1871519221000822. https://doi.org/10.1016/j.wombi.2021.05.003

Geoghegan, S., Stephens, L. C., Feemster, K. A., Drew, R. J., Eogan, M., & Butler, K. M. (2021). “This choice does not just affect me.” Attitudes of pregnant women toward COVID-19 vaccines: A mixed-methods study. Human Vaccines & Immunotherapeutics, 17(10), 3371–3376. https://doi.org/10.1080/21645515.2021.1924018

Glick, I., Kadish, E., & Rottenstreich, M. (2021). Management of Pregnancy in Women of Advanced Maternal Age: Improving Outcomes for Mother and Baby. International Journal of Women’s Health, Volume 13, 751–759. https://doi.org/10.2147/IJWH.S283216

Goldshtein, I., Steinberg, D. M., Kuint, J., Chodick, G., Segal, Y., Shapiro Ben David, S., & Ben-Tov, A. (2022). Association of BNT162b2 COVID-19 Vaccination During Pregnancy With Neonatal and Early Infant Outcomes. JAMA Pediatrics. https://doi.org/10.1001/jamapediatrics.2022.0001

Higgins, J. P. T. (2003). Measuring inconsistency in meta-analyses. BMJ, 327(7414), 557–560. https://doi.org/10.1136/bmj.327.7414.557

Hosokawa, Y., Okawa, S., Hori, A., Morisaki, N., Takahashi, Y., Fujiwara, T., Nakayama, S. F., Hamada, H., Satoh, T., & Tabuchi, T. (2022). The Prevalence of COVID-19 Vaccination and Vaccine Hesitancy in Pregnant Women: An Internet-based Cross-sectional Study in Japan. Journal of Epidemiology, JE20210458. https://doi.org/10.2188/jea.JE20210458

Huddleston, H. G., Jaswa, E. G., Lindquist, K. J., Kaing, A., Morris, J. R., Hariton, E., Corley, J., Hoskin, E., Gaw, S. L., & Cedars, M. I. (2022). COVID-19 vaccination patterns and attitudes among American pregnant individuals. American Journal of Obstetrics & Gynecology MFM, 4(1), 100507. https://doi.org/10.1016/j.ajogmf.2021.100507

Iacobucci, G. (2021). Covid-19 and pregnancy: Vaccine hesitancy and how to overcome it. BMJ (Clinical Research Ed.), 375, n2862. https://doi.org/10.1136/bmj.n2862

Khalil, A., von Dadelszen, P., Draycott, T., Ugwumadu, A., O’Brien, P., & Magee, L. (2020). Change in the Incidence of Stillbirth and Preterm Delivery During the COVID-19 Pandemic. JAMA. https://doi.org/10.1001/jama.2020.12746

Levy, A. T., Singh, S., Riley, L. E., & Prabhu, M. (2021). Acceptance of COVID-19 vaccination in pregnancy: A survey study. American Journal of Obstetrics & Gynecology MFM, 3(5), 100399. https://doi.org/10.1016/j.ajogmf.2021.100399

Lipkind, H. S., Vazquez-Benitez, G., DeSilva, M., Vesco, K. K., Ackerman-Banks, C., Zhu, J., Boyce, T. G., Daley, M. F., Fuller, C. C., Getahun, D., Irving, S. A., Jackson, L. A., Williams, J. T. B., Zerbo, O., McNeil, M. M., Olson, C. K., Weintraub, E., & Kharbanda, E. O. (2022). Receipt of COVID-19 Vaccine During Pregnancy and Preterm or Small-for-Gestational-Age at Birth—Eight Integrated Health Care Organizations, United States, December 15, 2020–July 22, 2021. MMWR. Morbidity and Mortality Weekly Report, 71(1), 26–30. https://doi.org/10.15585/mmwr.mm7101e1

Lokken, E. M., Huebner, E. M., Taylor, G. G., Hendrickson, S., Vanderhoeven, J., Kachikis, A., Coler, B., Walker, C. L., Sheng, J. S., Al-Haddad, B. J. S., McCartney, S. A., Kretzer, N. M., Resnick, R., Barnhart, N., Schulte, V., Bergam, B., Ma, K. K., Albright, C., Larios, V., … Washington State COVID-19 in Pregnancy Collaborative. (2021). Disease severity, pregnancy outcomes, and maternal deaths among pregnant patients with severe acute respiratory syndrome coronavirus 2 infection in Washington State. American Journal of Obstetrics and Gynecology, 225(1), 77.e1-77.e14. https://doi.org/10.1016/j.ajog.2020.12.1221

Mappa, I., Luviso, M., Distefano, F. A., Carbone, L., Maruotti, G. M., & Rizzo, G. (2021). Women perception of SARS-CoV-2 vaccination during pregnancy and subsequent maternal anxiety: A prospective observational study. The Journal of Maternal-Fetal & Neonatal Medicine, 1–4. https://doi.org/10.1080/14767058.2021.1910672

Mehraeen, E., Karimi, A., Barzegary, A., Vahedi, F., Afsahi, A. M., Dadras, O., Moradmand-Badie, B., Seyed Alinaghi, S. A., & Jahanfar, S. (2020). Predictors of mortality in patients with COVID-19–a systematic review. European Journal of Integrative Medicine, 40, 101226. https://doi.org/10.1016/j.eujim.2020.101226

Moher, D., Liberati, A., Tetzlaff, J., Altman, D. G., & The PRISMA Group. (2009). Preferred Reporting Items for Systematic Reviews and Meta-Analyses: The PRISMA Statement. PLoS Medicine, 6(7), e1000097. https://doi.org/10.1371/journal.pmed.1000097

Nikpour, M., Sepidarkish, M., Omidvar, S., & Firouzbakht, M. (2022). Global prevalence of acceptance of COVID-19 vaccines and associated factors in pregnant women: A systematic review and meta-analysis. Expert Review of Vaccines, 1–9. https://doi.org/10.1080/14760584.2022.2053677

Pinheiro, R. L., Areia, A. L., Mota Pinto, A., & Donato, H. (2019). Advanced Maternal Age: Adverse Outcomes of Pregnancy, A Meta-Analysis. Acta Médica Portuguesa, 32(3), 219. https://doi.org/10.20344/amp.11057

Pogue, K., Jensen, J. L., Stancil, C. K., Ferguson, D. G., Hughes, S. J., Mello, E. J., Burgess, R., Berges, B. K., Quaye, A., & Poole, B. D. (2020). Influences on Attitudes Regarding Potential COVID-19 Vaccination in the United States. Vaccines, 8(4), E582. https://doi.org/10.3390/vaccines8040582

Polack, F. P., Thomas, S. J., Kitchin, N., Absalon, J., Gurtman, A., Lockhart, S., Perez, J. L., Pérez Marc, G., Moreira, E. D., Zerbini, C., Bailey, R., Swanson, K. A., Roychoudhury, S., Koury, K., Li, P., Kalina, W. V., Cooper, D., Frenck, R. W., Hammitt, L. L., … C4591001 Clinical Trial Group. (2020). Safety and Efficacy of the BNT162b2 mRNA Covid-19 Vaccine. The New England Journal of Medicine, 383(27), 2603–2615. https://doi.org/10.1056/NEJMoa2034577

Rasmussen, S. A., & Jamieson, D. J. (2021). Pregnancy, Postpartum Care, and COVID-19 Vaccination in 2021. JAMA, 325(11), 1099–1100. https://doi.org/10.1001/jama.2021.1683

Rawal, S., Tackett, R. L., Stone, R. H., & Young, H. N. (2022). COVID-19 Vaccination among Pregnant People in the U.S.: A Systematic Review. American Journal of Obstetrics & Gynecology MFM, 100616. https://doi.org/10.1016/j.ajogmf.2022.100616

Razzaghi, H., Kahn, K. E., Black, C. L., Lindley, M. C., Jatlaoui, T. C., Fiebelkorn, A. P., Havers, F. P., D’Angelo, D. V., Cheung, A., Ruther, N. A., & Williams, W. W. (2020). Influenza and Tdap Vaccination Coverage Among Pregnant Women—United States, April 2020. MMWR. Morbidity and Mortality Weekly Report, 69(39), 1391–1397. https://doi.org/10.15585/mmwr.mm6939a2

Razzaghi, H., Meghani, M., Pingali, C., Crane, B., Naleway, A., Weintraub, E., Kenigsberg, T. A., Lamias, M. J., Irving, S. A., Kauffman, T. L., Vesco, K. K., Daley, M. F., DeSilva, M., Donahue, J., Getahun, D., Glenn, S., Hambidge, S. J., Jackson, L., Lipkind, H. S., … Patel, S. A. (2021). COVID-19 Vaccination Coverage Among Pregnant Women During Pregnancy—Eight Integrated Health Care Organizations, United States, December 14, 2020–May 8, 2021. MMWR. Morbidity and Mortality Weekly Report, 70(24), 895–899. https://doi.org/10.15585/mmwr.mm7024e2

Rottenstreich, M., Sela, H., Rotem, R., Kadish, E., Wiener□Well, Y., & Grisaru□Granovsky, S. (2022). Covid□19 vaccination during the third trimester of pregnancy: Rate of vaccination and maternal and neonatal outcomes, a multicentre retrospective cohort study. BJOG: An International Journal of Obstetrics & Gynaecology, 129(2), 248–255. https://doi.org/10.1111/1471-0528.16941

Santos, W. M. dos, Secoli, S. R., & Püschel, V. A. de A. (2018). The Joanna Briggs Institute approach for systematic reviews. Revista Latino-Americana de Enfermagem, 26(0). https://doi.org/10.1590/1518-8345.2885.3074

Sepandi, M., Taghdir, M., Alimohamadi, Y., Afrashteh, S., & Hosamirudsari, H. (2020). Factors Associated with Mortality in COVID-19 Patients: A Systematic Review and Meta-Analysis. Iranian Journal of Public Health. https://doi.org/10.18502/ijph.v49i7.3574

Shimabukuro, T. T., Kim, S. Y., Myers, T. R., Moro, P. L., Oduyebo, T., Panagiotakopoulos, L., Marquez, P. L., Olson, C. K., Liu, R., Chang, K. T., Ellington, S. R., Burkel, V. K., Smoots, A. N., Green, C. J., Licata, C., Zhang, B. C., Alimchandani, M., Mba-Jonas, A., Martin, S. W., … Meaney-Delman, D. M. (2021). Preliminary Findings of mRNA Covid-19 Vaccine Safety in Pregnant Persons. New England Journal of Medicine, 384(24), 2273–2282. https://doi.org/10.1056/NEJMoa2104983

Siegel, M. R., Lumbreras-Marquez, M. I., James, K., McBay, B. R., Gray, K. J., Schantz-Dunn, J., Diouf, K., & Goldfarb, I. T. (2021). Perceptions and Attitudes Towards COVID-19 Vaccination Amongst Pregnant and Postpartum Individuals [Preprint]. Obstetrics and Gynecology. https://doi.org/10.1101/2021.12.17.21267997

Skjefte, M., Ngirbabul, M., Akeju, O., Escudero, D., Hernandez-Diaz, S., Wyszynski, D. F., & Wu, J. W. (2021). COVID-19 vaccine acceptance among pregnant women and mothers of young children: Results of a survey in 16 countries. European Journal of Epidemiology, 36(2), 197–211. https://doi.org/10.1007/s10654-021-00728-6

Stock, S. J., Carruthers, J., Calvert, C., Denny, C., Donaghy, J., Goulding, A., Hopcroft, L. E. M., Hopkins, L., McLaughlin, T., Pan, J., Shi, T., Taylor, B., Agrawal, U., Auyeung, B., Katikireddi, S. V., McCowan, C., Murray, J., Simpson, C. R., Robertson, C., … Wood, R. (2022). SARS-CoV-2 infection and COVID-19 vaccination rates in pregnant women in Scotland. Nature Medicine, 28(3), 504–512. https://doi.org/10.1038/s41591-021-01666-2

Stuckelberger, S., Favre, G., Ceulemans, M., Nordeng, H., Gerbier, E., Lambelet, V., Stojanov, M., Winterfeld, U., Baud, D., Panchaud, A., & Pomar, L. (2021). SARS-CoV-2 Vaccine Willingness among Pregnant and Breastfeeding Women during the First Pandemic Wave: A Cross-Sectional Study in Switzerland. Viruses, 13(7), 1199. https://doi.org/10.3390/v13071199

Tao, L., Wang, R., Han, N., Liu, J., Yuan, C., Deng, L., Han, C., Sun, F., Liu, M., & Liu, J. (2021). Acceptance of a COVID-19 vaccine and associated factors among pregnant women in China: A multi-center cross-sectional study based on health belief model. Human Vaccines & Immunotherapeutics, 17(8), 2378–2388. https://doi.org/10.1080/21645515.2021.1892432

Taubman – Ben□Ari, O., Weiss, E., Abu□Sharkia, S., & Khalaf, E. (2022). A comparison of COVID-19 vaccination status among pregnant Israeli Jewish and Arab women and psychological distress among the Arab women. Nursing & Health Sciences, nhs.12929. https://doi.org/10.1111/nhs.12929

The Israeli Health Ministry. (2021). Recommendation that Pregnant Women Get Vaccinated with Frequent Exposure to People or with Chronic Medical Conditions. https://www.gov.il/en/departments/news/19012021-05

Townsel, C., Moniz, M. H., Wagner, A. L., Zikmund-Fisher, B. J., Hawley, S., Jiang, L., & Stout, M. J. (2021). COVID-19 vaccine hesitancy among reproductive-aged female tier 1A healthcare workers in a United States Medical Center. Journal of Perinatology, 41(10), 2549–2551. https://doi.org/10.1038/s41372-021-01173-9

Trostle, M. E., Limaye, M. A., Avtushka, V., Lighter, J. L., Penfield, C. A., & Roman, A. S. (2021). COVID-19 vaccination in pregnancy: Early experience from a single institution. American Journal of Obstetrics & Gynecology MFM, 3(6), 100464. https://doi.org/10.1016/j.ajogmf.2021.100464

UK Health Security Agency. (2021). COVID-19 vaccine surveillance report week 47. https://assets.publishing.service.gov.uk/government/uploads/system/uploads/attachment_data/file/1036047/Vaccine_surveillance_report_-_week_47.pdf

Vousden, N., Bunch, K., Morris, E., Simpson, N., Gale, C., O’Brien, P., Quigley, M., Brocklehurst, P., Kurinczuk, J. J., & Knight, M. (2021). The incidence, characteristics and outcomes of pregnant women hospitalized with symptomatic and asymptomatic SARS-CoV-2 infection in the UK from March to September 2020: A national cohort study using the UK Obstetric Surveillance System (UKOSS). PloS One, 16(5), e0251123. https://doi.org/10.1371/journal.pone.0251123

Wainstock, T., Yoles, I., Sergienko, R., & Sheiner, E. (2021). Prenatal maternal COVID-19 vaccination and pregnancy outcomes. Vaccine, 39(41), 6037–6040. https://doi.org/10.1016/j.vaccine.2021.09.012

Wallace, B. C., Schmid, C. H., Lau, J., & Trikalinos, T. A. (2009). Meta-Analyst: Software for meta-analysis of binary, continuous and diagnostic data. BMC Medical Research Methodology, 9(1), 80. https://doi.org/10.1186/1471-2288-9-80

Woodworth, K. R., Olsen, E. O., Neelam, V., Lewis, E. L., Galang, R. R., Oduyebo, T., Aveni, K., Yazdy, M. M., Harvey, E., Longcore, N. D., Barton, J., Fussman, C., Siebman, S., Lush, M., Patrick, P. H., Halai, U.-A., Valencia-Prado, M., Orkis, L., Sowunmi, S., … COVID-19 Pregnancy and Infant Linked Outcomes Team (PILOT). (2020). Birth and Infant Outcomes Following Laboratory-Confirmed SARS-CoV-2 Infection in Pregnancy—SET-NET, 16 Jurisdictions, March 29-October 14, 2020. MMWR. Morbidity and Mortality Weekly Report, 69(44), 1635–1640. https://doi.org/10.15585/mmwr.mm6944e2

Yanez, N. D., Weiss, N. S., Romand, J.-A., & Treggiari, M. M. (2020). COVID-19 mortality risk for older men and women. BMC Public Health, 20(1), 1742. https://doi.org/10.1186/s12889-020-09826-8

Zambrano, L. D., Ellington, S., Strid, P., Galang, R. R., Oduyebo, T., Tong, V. T., Woodworth, K. R., Nahabedian, J. F., Azziz-Baumgartner, E., Gilboa, S. M., Meaney-Delman, D., & CDC COVID-19 Response Pregnancy and Infant Linked Outcomes Team. (2020). Update: Characteristics of Symptomatic Women of Reproductive Age with Laboratory-Confirmed SARS-CoV-2 Infection by Pregnancy Status - United States, January 22-October 3, 2020. MMWR. Morbidity and Mortality Weekly Report, 69(44), 1641–1647. https://doi.org/10.15585/mmwr.mm6944e3

